# Polygenic scores for autism are associated with neurite density in adults and children from the general population

**DOI:** 10.1101/2024.04.10.24305539

**Authors:** Yuanjun Gu, Eva Maria-Stauffer, Saashi A. Bedford, APEX consortium, iPSYCH-autism consortium, Rafael Romero-Garcia, Jakob Grove, Anders D. Børglum, Hilary Martin, Simon Baron-Cohen, Richard A.I. Bethlehem, Varun Warrier

**Affiliations:** Department of Psychiatry, University of Cambridge, Cambridge, CB2 8AH; Autism Research Centre, Department of Psychiatry, University of Cambridge, Cambridge, CB2 8AH, UK; Department of Medical Physiology and Biophysics, Instituto de Biomedicina de Sevilla (IBiS), HUVR/CSIC/Universidad de Sevilla/CIBERSAM, ISCIII, 41013, Sevilla, Spain, 41013; The Lundbeck Foundation Initiative for Integrative Psychiatric Research, iPSYCH, Aarhus, 8210, Denmark; Center for Genomics and Personalized Medicine (CGPM), Aarhus University, Aarhus, 8000, Denmark; Department of Biomedicine (Human Genetics) and iSEQ Center, Aarhus University, Aarhus, 8000, Denmark; Bioinformatics Research Centre, Aarhus University, Aarhus, Denmark, 8000; Human Genetics Programme, Wellcome Sanger Institute, Wellcome Genome Campus, Hinxton, CB10 1SA, UK; Department of Psychology, University of Cambridge, Cambridge, CB2 3EB, UK

**Keywords:** Autism, polygenic scores, UK Biobank, ABCD, MRI, neuroimaging genetics

## Abstract

Genetic variants linked to autism are thought to change cognition and behaviour by altering the structure and function of the brain. Although a substantial body of literature has identified structural brain differences in autism, it is unknown whether autism-associated common genetic variants are linked to changes in cortical macro- and micro-structure. We investigated this using neuroimaging and genetic data from adults (UK Biobank, N = 31,748) and children (ABCD, N = 4,928). Using polygenic scores and genetic correlations we observe a robust negative association between common variants for autism and a magnetic resonance imaging derived phenotype for neurite density (intracellular volume fraction) in the general population. This result is consistent across both children and adults, in both the cortex and in white matter tracts, and confirmed using polygenic scores and genetic correlations. There were no sex differences in this association. Mendelian randomisation analyses provide no evidence for a causal relationship between autism and intracellular volume fraction, although this should be revisited using better powered instruments. Overall, this study provides evidence for shared common variant genetics between autism and cortical neurite density.

## Introduction

Autism is a highly polygenic, heritable set of neurodevelopmental conditions, characterised by restrictive repetitive behaviours and unusually narrow interests, sensory hyper- and hypo-sensitivities, and social-communication difficulties. Twin and familial recurrence studies indicate that heritability of autism is as high as 60-90% [1, 2], although single nucleotide polymorphism (SNP) heritability estimates are more modest, ranging from 11 - 50% [3, 4].

It is unclear how the polygenic likelihood for autism gives rise to the cognitive and behavioural outcomes that are collectively referred to as autism. In line with our theoretical understanding that autism emerges from changes in brain structure and function, genetic studies have demonstrated an enrichment for autism-related genes and genetic variants in early neurodevelopmental processes [3, 5–8]. Furthermore, studies have identified differences in cortical morphology (structural and diffusion phenotypes) and functioning in autistic compared to non-autistic individuals (e.g.,[9–11]) and among autistic individuals (e.g.,[12–17]). Studies have also identified associations between brain morphology and autistic traits in the general population (e.g., [18, 19]).

Previous work has demonstrated that at least a subset of autistic individuals shows differences in cortical volumes and surface area (SA) during development. For instance, longitudinal scans of autistic and non-autistic children have identified cortical enlargement in autistic children after the first year of life [20, 21]. Additionally, common genetic variants associated with typical differences in surface area and volume are enriched in genes identified from rare variant studies of neurodevelopmental conditions, including autism [22].

However, it is unclear if common and rare genetic variants associated with autism operate through similar biological and neural mechanisms. For instance, rare genetic variants implicated in autism are also associated with global developmental delay and intellectual disability [23]. In contrast, common genetic variants associated with autism contribute to higher cognitive ability [3, 24].

A small number of studies have investigated the neural correlates of common genetic variants associated with autism, indexed by polygenic scores (PGS). These studies demonstrated that PGS for autism is associated with global and regional alterations in cortical volume, cortical thickness (CT), and surface area (SA) [18, 25]. Of which, the PGS for autism association with CT is modified by age [26]. However, these studies were conducted in fewer than 2,500 individuals, which may be underpowered to detect small effect sizes using PGS for autism. A larger study in the UK Biobank demonstrated that PGS for autism is associated with multivariate differences in brain asymmetry [27].

Currently, there are no large-scale studies that systematically investigate how PGS for autism is associated with structural and diffusion derived brain imaging measures. We hypothesised that common genetic variants associated with autism may also be associated with a range of neuroimaging phenotypes, in both adults and children. In particular, we were interested in the role of autism-associated common genetic variants in cortical microstructure, given previous associations between cortical microstructure and genetic variation for schizophrenia and depression [28–30].

In parallel, studies have identified substantial sex differences in both brain structure [31–34] and the presentation and diagnosis of autism [35, 36]. Notably, even after accounting for social processes, there still is a male preponderance of autism diagnosis [36], suggesting that there may be biological factors that contribute to sex differences in autism. For instance, one study has demonstrated a shift in multivariate neuroanatomical patterns in autistic individuals towards that typically observed in males [31]. Subsequently, we were also interested in investigating if sex differences in autism are reflected in the sex differential effects of PGS for autism on brain structure.

Here, we address these questions using the largest homogenous imaging genetics dataset available from the UK Biobank (N=31,748) and ABCD (N=4,928). We investigated global and regional association between genetic likelihood for autism and brain structural changes in the general population, as well as potential sex differences in this association **(Figure 1)**. We studied three macrostructural MRI-derived phenotypes and two microstructure phenotypes both globally as well as 180, bilaterally averaged cortical regions based on the Glasser parcellation. We focus on these five phenotypes, as these five are highly correlated with other cortical phenotypes, index five different latent traits, and have relatively high heritability compared to several other measurable phenotypes [22]. Follow-up analyses such as genetic correlation and Mendelian randomisation were conducted to investigate the robustness of the results and causality, respectively. Finally, to contextualise the results, we also ran enrichment analyses for the autism PGS association with known functional networks [37] and cytoarchitectonic classes [38].

**Fig. 1:**
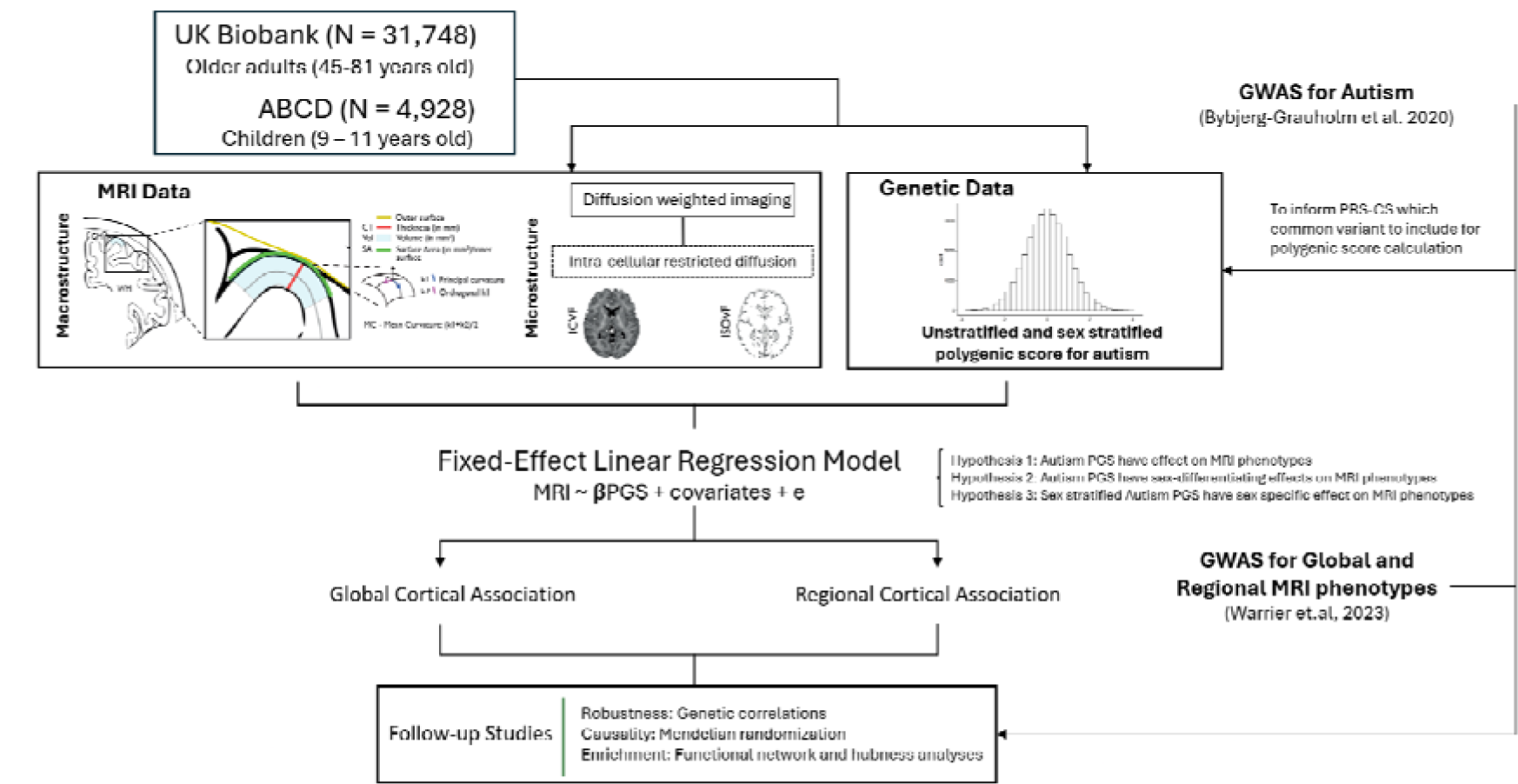
Schematic summary. We generated polygenic scores for autism in two datasets - UK Biobank and ABCD and investigated the associations between the polygenic scores and five brain structural phenotypes globally as well as in 180 regions. We fitted sex-stratified and unstratified models. We confirmed the robustness of the results using genetic correlations and assessed causality using Mendelian randomization. Finally, to contextualise the results, we investigated enrichment of the associations in cortical networks.

## Methods

### Participants

This study is based on data from two independent cohorts, the UK Biobank (UKB) [39] and the Adolescent Brain Cognitive Development (ABCD) database [40]. In both cohorts, we excluded participants with incomplete MRI and genotype data and focused on participants with genetically inferred European ancestry. This generated two subgroups, consisting of N = 31,748 UKB participants and N = 4,928 ABCD participants. Details of age, sample size, and sex are summarised in **Supplementary Table (ST) 1**. Ethical approval was obtained from the Human Biology Research Ethics Committee, University of Cambridge (Cambridge, UK). Informed consent was provided by all participants.

### Acquisition, preprocessing and quality control for imaging data

For MRI data, minimally preprocessed T1 and T2-FLAIR weighted data were downloaded from UKB (application 20904) and ABCD through their respective data repository, and further processed using FreeSurfer version (Version 6.0.1) ([41]) for cortical reconstruction [22].

Then, the data was parcellated using the Human Connectome Project parcellation (i.e. Glasser parcellation) [42] using surface-to-surface mapping to align the diffusion and T1-weighted imaging. The recon-all reconstruction used both T1- and T2-weighted images when both were available, and all subsequent statistical analyses included a covariate for the type of reconstruction (i.e., T1 only or T1-T2 combined). Reconstruction quality was evaluated using the Euler index [43] and was included as a covariate in subsequent statistical analyses. Macrostructural MRI-derived phenotypes, including total surface area of the cortex (SA), average cortical thickness (CT), and total mean curvature (MC) were extracted and standardised for further analysis.

Reconstruction of Neurite orientation dispersion and density imaging (NODDI) was generated by the Accelerated Microstructure Imaging via Convex Optimization (AMICO) pipeline [44] and was then aligned to the Glasser parcellation. NODDI can measure three types of microstructural environment in the human brain. In this study, we used two of them: a measure of the density neurites including axon and dendrites by measuring the extent of intracellular diffusion (Intracellular Volume Fraction or ICVF) and a measure of isotropic volume, typically thought to represent free-water or the cerebrospinal fluid (Isotropic Volume Fraction or ISOVF). Note ICVF is also called Neurite Density Index (NDI).

Global values of the MRI-derived phenotypes were obtained by averaging or summing all values from the 360 regions. Regional values of MRI-derived phenotypes were bilaterally averaged, resulting in 180 regional values. Outliers were removed by excluding individuals with more than 5 standard deviations (SD) or median absolute deviations beyond the mean and median respectively.

In addition, as previously conducted [29], probabilistic tractography was used to calculate ICVF values at each of 15 major white matter tracts defined using AutoPtx [45].

### Acquisition and quality control for genetic data

Details of the genome-wide genotype data used for this study including processing, imputation, and quality control can be found in detail elsewhere for UKB [46] and ABCD [22, 47].

Briefly, in both datasets, we excluded participants who were not of genetically inferred European ancestry based on self-reported data and genetic principal component-based clustering. From the group of included participants, we further excluded individuals with less than 95% genotyping rate, who had a mismatch between genetic and reported sex, and who had excess genetic heterozygosity. Additionally, we also excluded any individuals who were more than ± 5 standard deviations from the means of the first two genetic principal components to minimise the impact of fine-scale population stratification on the analyses.

In both UKB and ABCD, we included genotyped and imputed SNPs with a minor allele frequency > 0.1%, Hardy-Weinberg equilibrium (P > 1 x 10^-6^), genotyping rate > 95%, and, for imputed SNPs, imputation quality R^2^ > 0.4.

### Polygenic scores

Unstratified and sex-stratified polygenic scores (PGS) for autism were calculated for UKB and ABCD using PRS-CS [48], based on effect sizes of autosomal SNPs provided by autism GWAS summary statistics from the iPSYCH cohort [49]. The iPSYCH GWAS consists of 19,870 autistic participants (15,025 males) and 39,078 non-autistic individuals (19,763 males). All PGS were standardised to have a mean of zero and a standard deviation of one. We used the iPSYCH GWAS compared to the publicly available Grove et al., 2019 GWAS [3] because the iPSYCH had a larger sample size, better statistical power (mean X^2^ = 1.23 for iPSYCH GWAS and 1.2 for Grove et al., 2019), is unlikely to be affected by participation bias, and because we had access to sex-stratified GWAS data from the same sample.

### Statistical analyses

All statistical analyses between PGS for autism and MRI-derived phenotypes were conducted in R (Version 4.3.1).

We first investigated whether PGS for autism is associated with variation in MRI-derived global and regional phenotypes (Hypothesis 1, Equations 1 and 2). To identify the effect of PGS for autism on region-specific effects, global values of MRI-derived phenotypes of interest were included as covariates in conditional analyses (Equation 3, 5, 7 and 9).

Given that the mean scores of the MRI-derived phenotypes differed between sexes (**ST 2**), for sex differential effects, we investigated if PGS for autism is differentially associated with variation in MRI-derived phenotypes by sex by including a sex x PGS interaction term into the model (Hypothesis 2). We tested this across both global and regional phenotypes (Equations 4 - 5). The genetic correlation between the two sex-stratified autism GWAS, although high, was significantly less than 1. Given this, we also investigated whether sex-stratified PGS for autism will have sex-specific effects on MRI-derived phenotypes in males and females separately both globally and regionally (Hypothesis 3, Equations 6 - 9).

For all regressions we included age, age^2^, the first 10 genetic principal components, genotype sequencing batch, scanning site number, Euler index, framewise displacement [22], and T1-T2 scan status. These covariates were added to minimise the effect of confounding variables, based on availability within the cohort database. For Hypothesis 1 we further included sex, sex x age, and sex x age^2^ as covariates.

All genetic and imaging data were standardised before and after sex stratification. All continuous technical covariates, such as Euler Index were also similarly standardised. For all associations with regional MRI-derived phenotypes, the p values were adjusted for multiple testing for the total number of Glasser parcellation regions for each hemisphere (n = 180) using Benjamini-Hochberg false discovery rate (FDR) [50]. We used an FDR-corrected p value ≤ 0.05 as the statistical significance threshold.

### Equations underlying the Linear Regression Models

**Hypothesis 1:** Autism PGS is associated with variations in MRI-derived phenotypes:

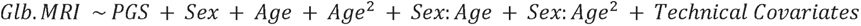

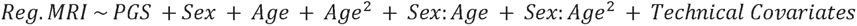

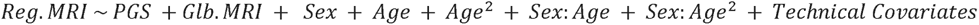

**Hypothesis 2**: Autism PGS has sex-differential association with MRI-derived phenotypes:

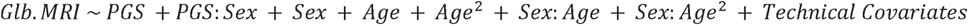

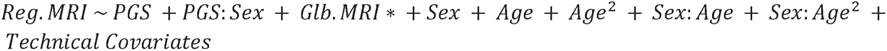

**Hypothesis 3:** Sex stratified autism PGS have sex-specific associations with MRI-derived phenotypes:

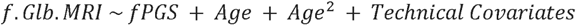

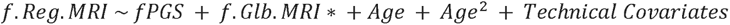

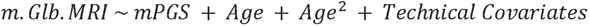

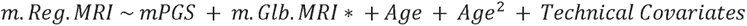

Note: MRI indicates the MRI-derived phenotype of interest. Prefix Glb. indicates global MRI-derived phenotype value; prefix Reg. indicates Regional MRI-derived phenotype; prefix f. indicates female-stratified values; prefix m. indicates male-stratified values. For equations 5, 7 and 9, we ran separately with and without the global MRI-derived phenotypes as covariates. This is indicated by Glb.MRI*.

### Bidirectional Mendelian randomization analyses and genetic correlations

We investigated the causal relationship between autism and the global and regional imaging phenotypes using two-sample bidirectional Mendelian randomization (MR) analyses, using the ‘TwoSampleMR’ (Version 0.5.7) and ‘MRPRESSO’ (Version 1.0) packages in R. Specifically, we were interested in understanding if (i) autism (exposure) causes changes in brain structure indexed by MRI-derived phenotypes (outcome) and/or (ii) changes in brain structure indexed by MRI-derived phenotypes (outcome) causes autism (outcome). For autism, we used SNPs from the unstratified autism GWAS from iPSYCH. For MRI-derived phenotypes, we used the meta-analysed GWAS of ABCD and UKB [22]. For all GWAS of exposure phenotype, we included only SNP genetic instruments that reached genome-wide significance (p < 5L×L10^−8^), after clumping at 10,000 kb distance and an LD r^2^ of 0.001.

We fitted different MR models, including inverse variance weighted (IVW) MR, median weighted (majority valid), MR Egger (accounts for pleiotropy,) and MR PRESSO (detects and excludes outliers in the instrument) to test the robustness of the results. We conducted further sensitivity analyses, such as testing for heterogeneity and horizontal pleiotropy, and leave-one-out to interrogate the validity of the results.

Genetic correlations between autism and global and regional MRI-derived phenotypes were estimated through LDSC (Version 1.0.1) [51], using Autism GWAS summary statistics from the iPSYCH cohort [49] and meta-analysed GWAS summary statistics of each MRI-derived phenotypes from the combined ABCD and UKB cohorts [22]. The LD score used is provided by the North-West European population from the 1000 Genomes project [52]. For genetic correlation between autism and regional MRI-derived phenotypes, only regions that have shown significant association in equation 2 are included in the analysis.

All results were corrected for multiple corrections using Benjamini Hochberg False Discovery Rate (FDR) corrected p value ≤ 0.05.

### Network and enrichment analyses

We ran a series of investigations to understand the links between brain networks and PGS for autism. Conceptualising the brain as a connected network, we generated hubness scores for each region by calculating both mean phenotypic (structural connectivity hubness) and genetic (genetic hubness) correlation between each cortical region and every other cortical region. Structural connectivity hubs were calculated using phenotypic data separately in the UKB and ABCD. For genetic hubness we used the genetic correlation values from the meta-analysed GWAS of ABCD and UK Biobank [22]. We investigated the correlation between hubness scores and PGS association across regions.

We further investigated if the association of PGS with the regional phenotypes were enriched in cortical networks identified from intrinsic functional connectivity (Yeo-Kreinen networks [37]) and cytoarchitectonic classes (Mesulam classes [38]). Enrichment analyses were conducted using 1,000 “spin” permutation testing [53], which accounts for the spatial autocorrelation among regions. Results were FDR-corrected for each MRI-derived phenotype and cortical atlas.

### Sex-stratified GWAS of global imaging phenotypes

To investigate potential sex differences, we conducted GWAS of global SA, CT, MC, ICVF and ISOVF in the UK Biobank separately in males (N = 14,957) and females (N = 16,833). We followed the same pipeline as used before [22] and genetic quality control is detailed earlier in the section “Acquisition and quality control of genetic data”. All phenotypes were standardised, and we included the same covariates included in the PGS analyses to test Hypothesis 1. GWAS was conducted using fastGWA [54] which uses a linear mixed effects model to account for fine-scale population stratification and relatedness among participants. We used the sex-stratified GWAS for genetic correlation analyses with the sex-stratified autism GWAS to investigate sex-specific effects.

### Data availability

This study used imaging, genetic and demographic data collected from the UK Biobank Resource under application number 20904, and ABCD. Data are available by request to registered and approved researchers on their corresponding platforms.

GWAS summary statistics and cortical morphology data are available through request from the CAM:IDE Data Access Portal (https://portal.ide-cam.org.uk/overview/483)[22]. iPSYCH-based GWAS data for autism [49] can be requested from Jakob Grove and Anders Børglum.

### Code availability

All analyses were written in R (Version 4.3.1). Mendelian Randomisation was performed using R package TwoSampleMR [55, 56] (https://mrcieu.github.io/TwoSampleMR/) and MRPRESSO [57] (https://github.com/rondolab/MR-PRESSO).

The pipelines used to generate processed MRI-derived phenotypes are available through GitHub (UKB, https://github.com/ucam-department-of-psychiatry/UKB; ABCD, https://github.com/ucam-department-of-psychiatry/ABCD. PGS was generated using the code provided by PRScs using “--phi 1e-2”: https://github.com/getian107/PRScs. Genetic correlations were conducted in line with the code provided by the software developers: https://github.com/bulik/ldsc. Details of how glasser regions were mapped onto the networks are provided here: https://github.com/ucam-department-of-psychiatry/maps_and_parcs.

All other codes used for this study are available at https://github.com/yg330/autism.img-pgs.

## Results

### Autism polygenic scores are associated with reduced global cortical ICVF

We first investigated the association between PGS for autism and five global (standardised summed or averaged) structural or diffusion MRI-derived phenotypes in both the UK Biobank and ABCD. These include surface area (SA), cortical thickness (CT), mean curvature (MC), Intracellular Volume Fraction (ICVF, also called Neurite Density Index or NDI), and Isotropic Volume Fraction (ISOVF). We chose these five phenotypes as they are highly correlated with other structural and diffusion phenotypes representing five cortical latent traits, have low correlations between each other, and have higher SNP heritability compared to other highly correlated phenotypes [22]. In the UKB, PGS for autism was significantly associated with lower SA (Incremental variance explained i.e., Inc R^2^: 1.23e-04) and ICVF (Inc R^2^: 1.23e-04), and increased CT (Inc R^2^: 2.10e-04) after FDR correction (**Figure 2A** and **ST 3**). In the younger ABCD cohort, higher PGS for autism was nominally significantly associated with lower ICVF (Inc R^2^: 7.31e-04) and significantly with higher ISOVF (Inc R^2^: 1.26e-03) (**ST 3**). We further confirmed the robustness of these observations through genetic correlation, where we observed a significant negative genetic correlation between ICVF and autism (r_g_ = -0.22, s.e.m = 0.07, p = 1.80e-3) (**Figure 2B** and **ST 4**).

**Fig 2.**
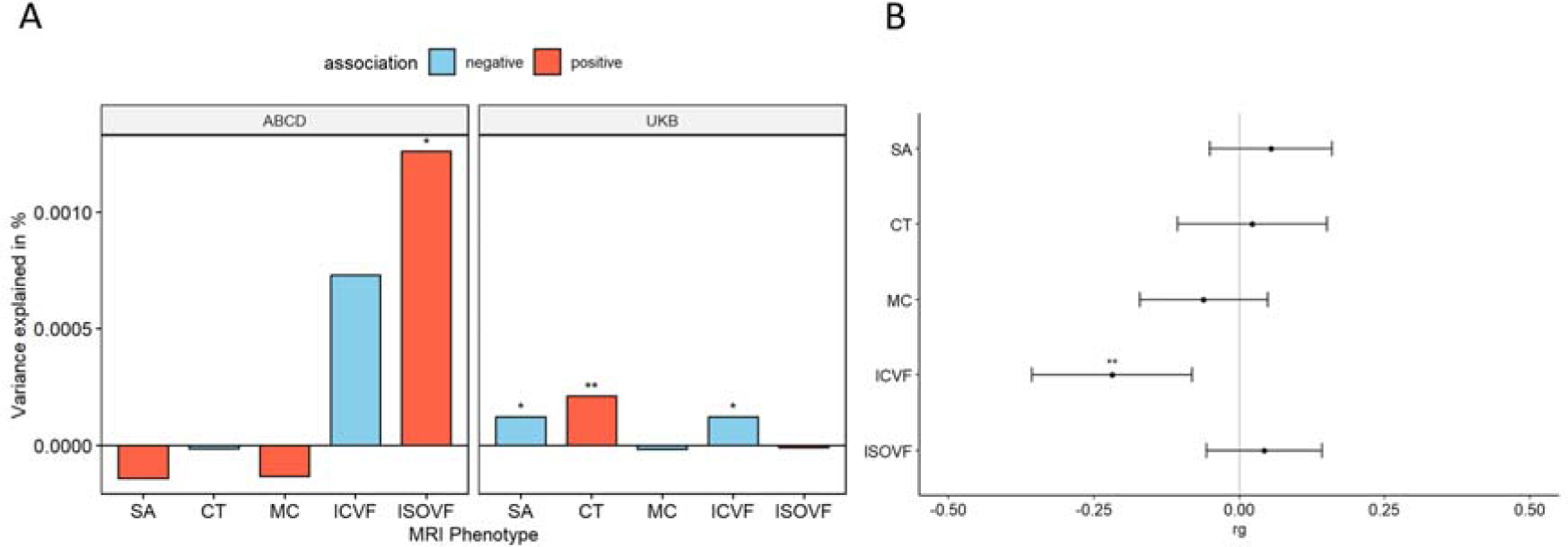
Statistical and Genetic correlation between polygenic scores for autism and global MRI-derived phenotypes. **A** Percentage of variance explained by polygenic score for autism (PGS) for different global cortical MRI-derived phenotypes under cohorts. SA surface area, CT cortical thickness, MC mean curvature, ICVF intracellular volume fraction or neurite density index, ISOVF isotropic volume fraction. Blue bar indicates negative associations, and the red bar indicates positive associations. * p <= 0.05, ** p <= 0.01. **B** Genetic correlation (rg) between autism and MRI-derived phenotype GWAS. Whiskers indicate 95% confidence intervals. Asterisks indicate p values after FDR<5% correction, with code sign the same as A.

### PGS for autism are negatively associated with regional cortical ICVF

We next investigated whether PGS for autism are associated with regional variation in the same five MRI-derived phenotypes. In the UKB, we identified a negative association between PGS for autism and ICVF in nine regions (Inc.R^2^: 2.25e-4 to 5.72e-4) and with SA in 24 regions (Inc.R^2^: 1.53e-4 to 3.60e-4) after FDR correction (**Figure 3A**). We did not identify a significant association with any other regional phenotypes (**ST 5**).

**Figure 3.**
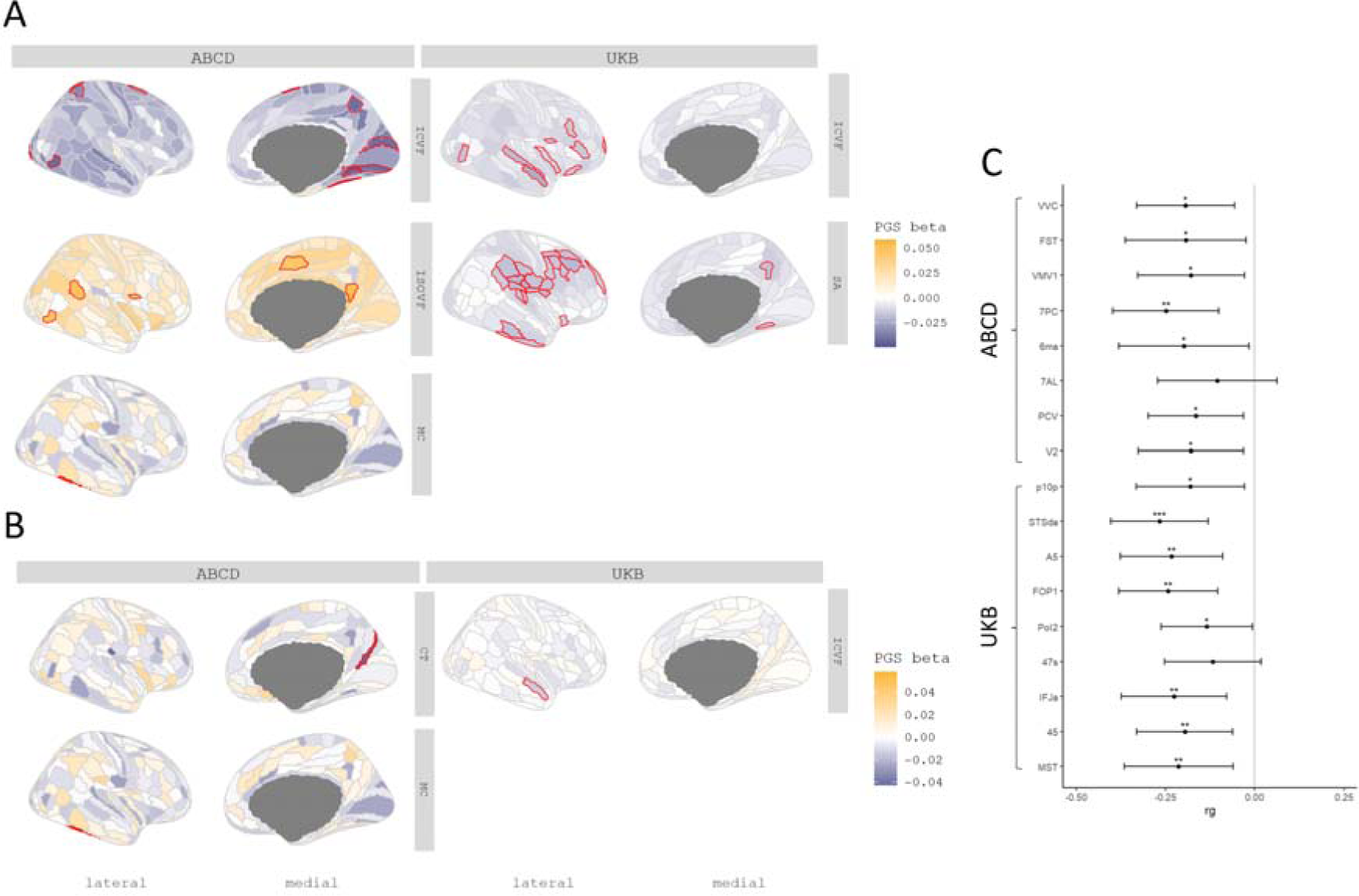
Statistical and Genetic correlation between polygenic scores for autism and regional MRI-derived phenotypes. **A** Cortical map of regional associations between PGS for autism and cortical MRI-derived phenotypes. Red outline means the effect of PGS for autism reached statistical significance at FDR <=0.05. **B** Cortical map of regional associations between PGS for autism and cortical MRI-derived phenotypes after global phenotype correction. Red outline means the effect of PGS for autism reached statistical significance at FDR <=0.05. For brevity, we have displayed only regional association maps for phenotypes where there was at least one significant regional association. **C** Genetic correlation between autism and regional ICVF in regions with statistically significant associations with the PGS for autism. Asterisks indicate p values after FDR correction: * p <= 0.05, ** p <= 0.01, *** p <= 0.001.

In ABCD, despite the smaller sample size, ICVF was significantly negatively associated with PGS for autism in eight regions (Inc.R^2^: 1.64e-3 to 2.11e-3) (**Figure 3B**), although these regions differed from the significantly associated regions in the UKB (**ST 6**). In the UKB, the significant associations with ICVF were primarily in the frontal and temporal lobes whereas in ABCD, the significant associations were in the occipital lobe. In addition, PGS for autism was positively associated with MC (Inc.R^2^: 3.20e-3) in the lateral temporal region, and ISOVF in five regions (Inc.R^2^: 1.96e-3 to 2.36e-3).

Supporting the robustness of the associations with ICVF, autism had significant and negative genetic correlations with ICVF in 15 of the 17 regions that were significantly associated with autism PGS (**ST 4** and **Figure 3C**). In contrast, we did not identify any significant associations between autism and SA, MC and ISOVF. Together, these analyses identify robust shared genetics between autism and ICVF using two different methods and in two different cohorts.

To account for the effect of the global phenotypes, we re-ran the analyses after correcting for the global phenotypes. In these conditional analyses, ICVF in one region, STSda (Inc.R^2^: 2.26e-4) located in the superior temporal gyrus, and MC in TE2P, located in the fusiform gyrus (Inc.R^2^: 3.19e-3), remained significantly associated with PGS for autism. We additionally found one region in CT named DVT, located in the posterior cingulate, which showed a significant association with PGS in autism (**ST 7** and **ST 8**).

### Autism PGS is more negatively associated with ICVF in cortical hubs

We recognise that cortical regions are not truly independent but may be organised as networks based on structural or genetic similarity, or functional co-activation [37], or cellular similarity [38]. For example, CT in regions that co-vary or are connected with several other regions (termed hubs) were more likely to be altered in autism [58]. Investigating this hypothesis, we observe a significant correlation between genetic hubness and PGS association with autism for ICVF in both the UKB and ABCD, and CT in ABCD (**ST 9, Figure 4**). Further supporting this, and in line with Cheverud’s conjecture, we identified a significant correlation between structural connectivity and PGS association for ICVF in both ABCD and UKB. We also identified a significant association between structural connectivity and autism PGS association with CT in the ABCD and ISOVF in the UKB.

**Fig 4.**
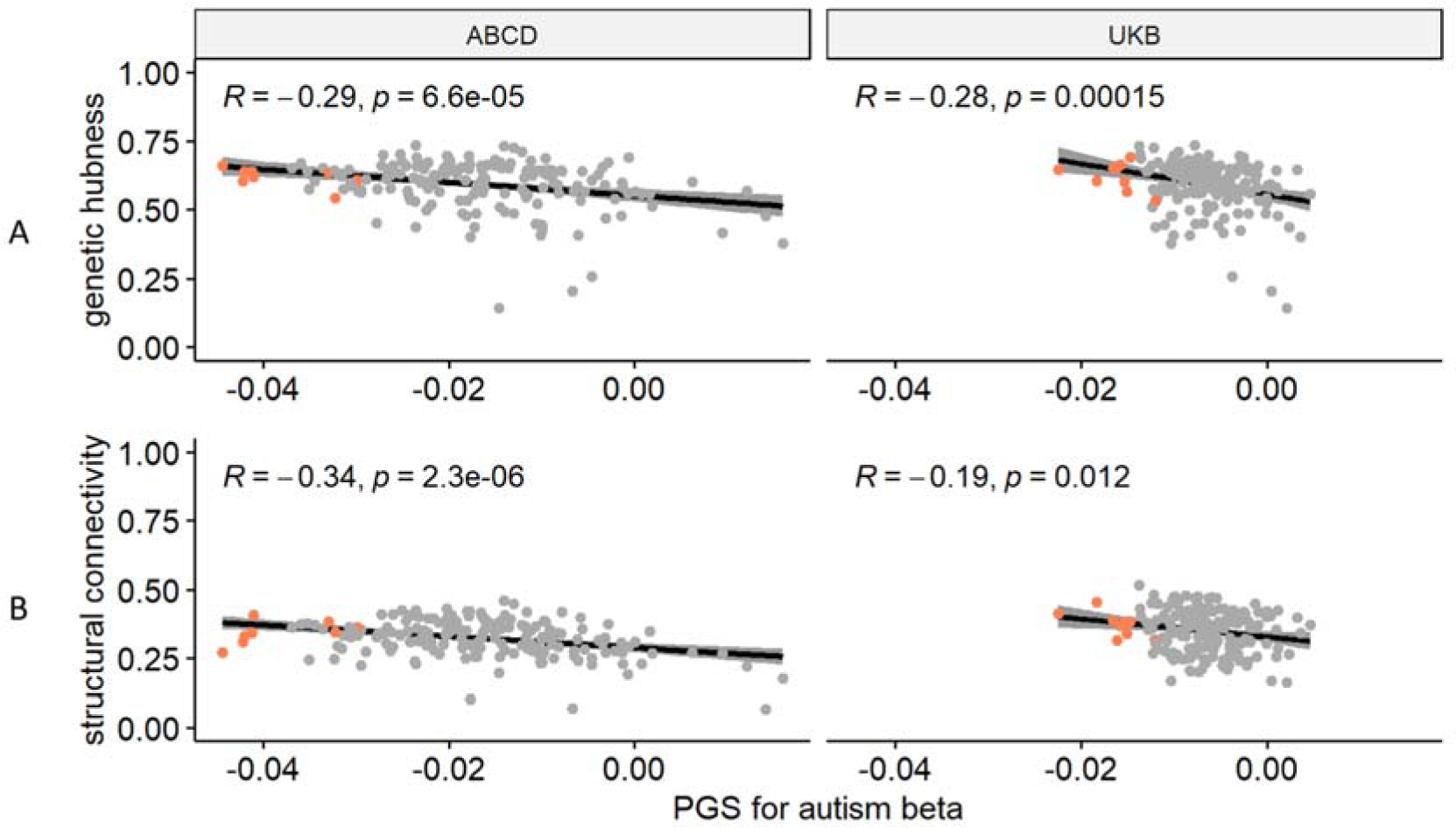
Enrichment Analysis: **A** Correlation plot between genetic hubness and autism PGS association with ICVF in the ABCD and UK Biobank cohorts. **B** Correlation plot between structural connectivity and autism PGS association with ICVF in the ABCD and UK Biobank cohorts. Each point represents a region and regions where there is a significant association between the phenotype and autism PGS are coloured in red. R is the Pearson correlation coefficient. p is the p value before correction for multiple testing, all p-value remained significant (<0.05) after FDR correction.

In addition to the significant association between hubness and ICVF, regions with statistically significant associations with autism PGS also on average had higher hubness (**Figure 4, Supplementary Figures 1 and 2**). We further considered if there is an association of the PGS effects in known functional networks (Yeo) or cytoarchitectural classes of the cortex (Mesulam). After spin permutation correction, PGS associations for MC were enriched in the idiotypic cortex (Mesulam class) in both ABCD and UKB (**ST 10**). We did not identify any other robust enrichments across both ABCD and UKB.

### PGS for autism are negatively associated with ICVF in white matter tracts

As PGS for autism consistently showed significant negative associations with cortical ICVF, we were interested in investigating if similar effects would be observed for 27 white matter tracts. We investigated this only in the UKB due to data availability. PGS for autism was significantly negatively associated with ICVF in 22 out of 27 ICVF white matter tracts (Inc R2 from -1.77e-05 to 2.15e-04) (**ST 11 and Figure 5**). The variance in white matter tracts ICVF explained by the autism PGS was similar to that of regional cortical ICVF in the UKB. However, in contrast to associations with cortical ICVF, we did not observe significant genetic correlations between autism and ICVF in white matter tracts (**ST 4**).

**Fig 5.**
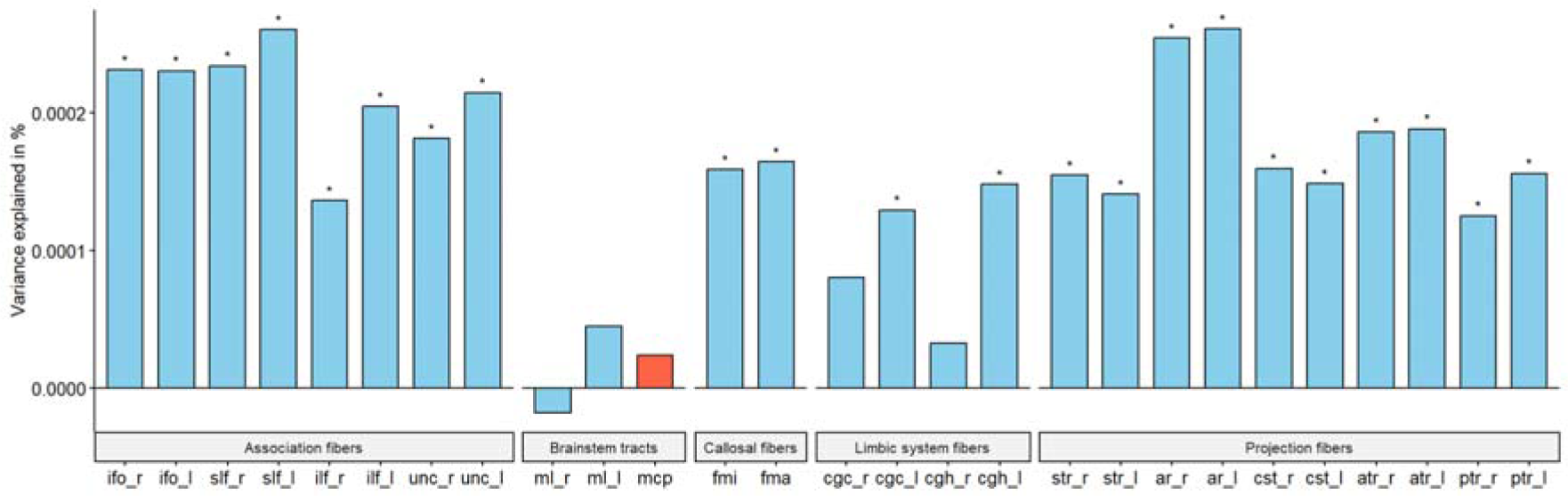
White tract association. Percentage of variance explained by polygenic score for autism (PGS) for 15 major white matter tracts for ICVF in UKB. Blue bar indicates negative associations, and the red bar indicates positive associations. * p <= 0.05. The acronyms for the 15 major white matter tracts are mcp = middle cerebellar peduncle, ml = medial lemniscus, cst = corticospinal tract, ar = acoustic radiation, atr = anterior thalamic radiation, str = superior thalamic radiation, pts = posterior thalamic radiation, slf = superior longitudinal fasciculus, ilf = inferior longitudinal fasciculus, ifo = inferior fronto-occipital fasciculus, unc = uncinate fasciculus, cgc = cingulate gyrus part of cingulum, cgh = parahippocampal part of cingulum, fmi = forceps minor, and fma = forceps major. Suffix _r means it is estimated from the right hemisphere and _l means it is estimated from the left hemisphere.

**Figure 6:**
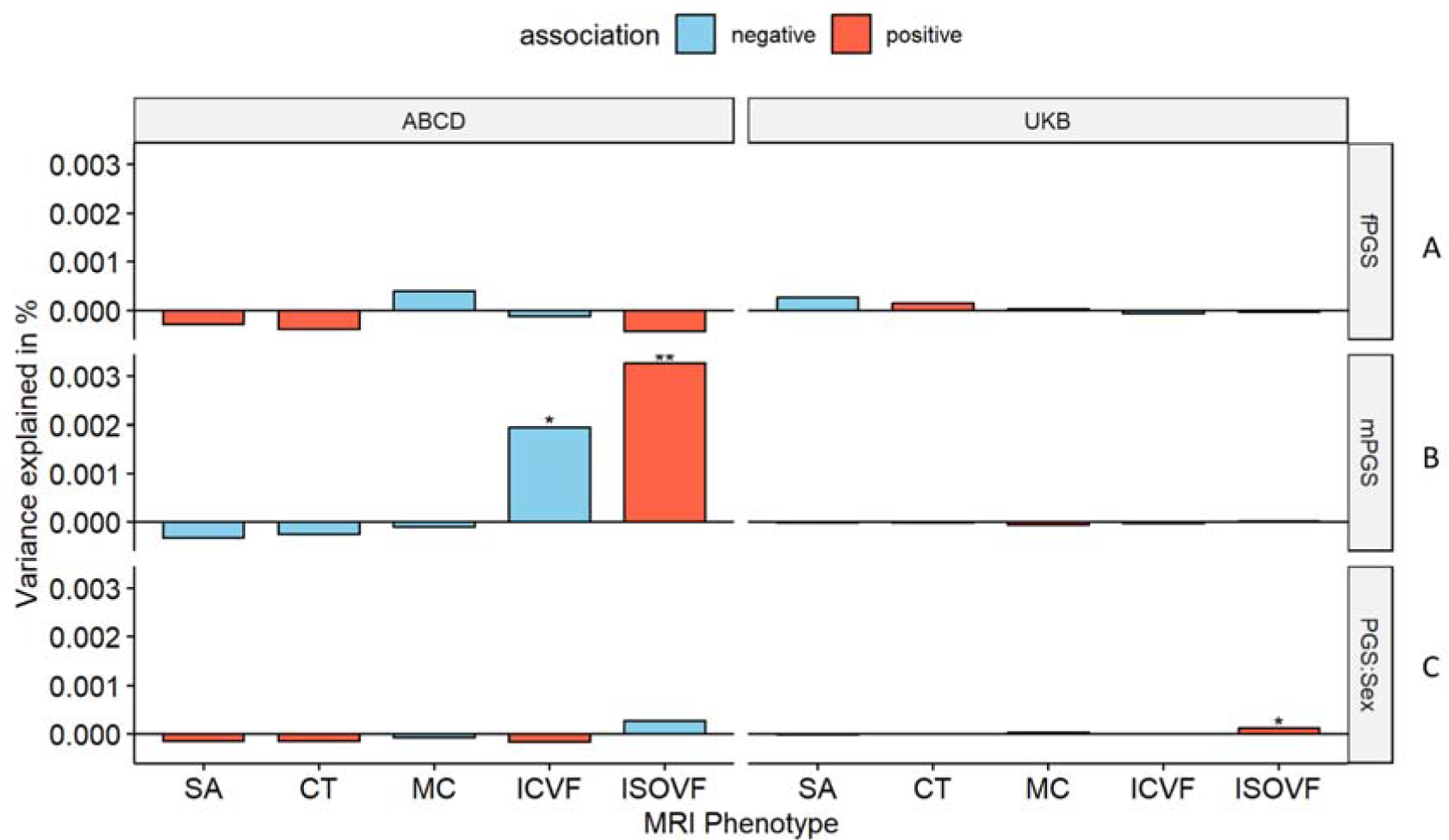
Percentage of variance explained by sex-specific and sex-differentiating PGS for autism. **A and B** Percentage of variance explained by sex stratified autism PGS for the global phenotypes in the equivalent sexes in ABCD and UKB. **C** Percentage variance explained by the sex interaction analysis. Asterisks indicate P values after FDR correction: * P <= 0.05, ** P <= 0.01. mPGS = polygenic scores from the males-only GWAS, fPGS = polygenic scores from the females-only GWAS, and PGS:Sex = the interaction between sex and PGS. SA = surface area; CT = cortical thickness, MC = mean curvature, ICVF = intracellular volume fraction.

### No evidence for causal relationships between autism and MRI-derived phenotypes

Since there were significant genetic correlations between PGS for autism and global and regional MRI-derived phenotypes, we additionally investigated whether there is a casual relationship between autism and these MRI-derived phenotypes using four different Mendelian randomisation methods. Specifically, we focussed on the phenotypes (global and regional) where we observed a significant association with PGS, hypothesising that causal relationships will lead to shared genetics. Additionally, for regional phenotypes, we excluded any phenotype with fewer than three genome-wide significant SNPs after harmonisation.

For global MRI-derived phenotypes, after adjusting for multiple correction based on MRI-derived phenotype tested, we found no significant evidence for causal effect of MRI-derived phenotypes on autism, nor causal effect of autism on MRI-derived phenotypes across four MR methods tested (**ST 12** and **13**). Similar results were also seen for autism and regional MRI-derived phenotypes after multiple testing corrections (**ST 14** and **15**).

### Limited evidence for sex differential effects of autism PGS on brain structure

Given that more males are diagnosed as autistic compared to females, we investigated if some of the sex differences in both the manifestation and diagnosis of autism are attributable to sex-differential effects in the cortex. The sex-stratified GWAS of autism from iPSYCH had high genetic correlation (r_g_ = 0.80, s.e.m = 0.08, p = 2.22e-20), suggesting that the common variant effects for autism are largely similar between sexes.

We first investigated if the effects of autism PGS on MRI-derived phenotypes are statistically different between sexes by including a PGS x sex interaction term. For the global phenotypes, we found a significant sex by PGS interaction for ISOVF in UKB (**ST 3**), but this was not significant in ABCD (**ST 3**) nor generalisable to regional ISOVF phenotypes (**ST 16** and **ST 17**). In ABCD, we found regional sex differential associations for CT in three regions only after global phenotype correction, although this was not significant in the UKB (**ST 18** and **ST 19**). Overall, we did not find robust evidence to suggest sex differential effects of the autism PGS on cortical MRI-derived phenotypes.

The GWAS for autism in males and females showed high correlation (r_g_ = 0.80, s.e.m = 0.08, p = 2.22e-20), but the correlation was still significantly less than 1. This suggests the possibility of sex-specific genetic influences on autism. To further investigate potential sex differences, we used the sex-stratified autism GWAS to generate sex-specific polygenic risk scores for autism. We then examined the effects of these sex-specific scores in individuals of the same sex in the UK Biobank and ABCD globally (**ST 3**) and regionally (**ST 20** to **ST 27**). In UKB, we did not observe any sex-specific effect of PGS for autism. In ABCD, we observed significant male-specific autism PGS effects on ICVF and ISOVF in males (**ST 3**). Regionally, we found no male-specific, nor female-specific associations in UKB. In ABCD, we found male-specific regional effects for ICVF in one region (PCV in the precuneus) and ISOVF in nine regions. Additionally, after global phenotype correction, we identified male-specific regional effects for ISOVF in three regions. We did not find any significant genetic correlation between the sex-stratified autism GWAS and sex-stratified global MRI-derived phenotypes after multiple testing correction (**ST 4**).

## Discussion

There has been a longstanding interest in how the genetic likelihood for autism manifests in the brain. Here, we investigate this by interrogating the correlates of common genetic variants for autism and MRI-derived brain phenotypes. In the largest sample to date, we systematically investigate the relationship between PGS for autism and MRI-derived phenotypes, beyond CT and SA [18, 26], and also investigate how sex and sex differences modify these relationships.

We found widespread negative associations between PGS for autism and ICVF, a measure of neurite density. ICVF, measured using the NODDI orientation sequence, is thought to be a measure of the density of neurites (axons and dendrites) in the brain, which has been supported by histological studies in humans [59] and mice [60]. This association was observed in both adults and children and validated using two different methods: polygenic scores and genetic correlations. This age-invariant association is noteworthy as neurite density changes non-linearly with age: it increases rapidly during childhood and decreases in later adulthood [61, 62]. One explanation of this robust association across ages is that the genetic likelihood for autism, alters the development of neurite density across development. Although Mendelian Randomisation did not identify a causal relationship, this could be due to the reduced statistical power of the instruments used. This association between autism and reduced neurite density is supported by previous case-control imaging studies [63–65] and mice studies focusing on autism-associated genes [66, 67]. These findings are paralleled by early postmortem brain studies in autism, which have identified cortical disorganisation [68] and reduced mini-columns in autism [69]. However, these findings have typically been conducted in small cohorts, and have not been consistently replicated. Furthermore, it is difficult to map microscale changes in neural cytoarchitecture, identified from postmortem studies, to relatively gross changes captured by MRI.

The association with reduced ICVF, at least globally, is not specific to autism, but also associated with many other traits and conditions, such as sleep, seizure, psychosis, schizophrenia, and Parkinson’s disease [29, 70, 71]. For instance, previous work has identified that PGS for schizophrenia is associated with reduction in global and regional ICVF [29]. However, the effects of the autism PGS on ICVF were only modestly correlated with the effects of schizophrenia PGS on ICVF (r = 0.33, 95% CI = 0.19 - 0.45), suggesting that although both autism and schizophrenia PGS are associated with reduction in ICVF, their effects vary. Notably, this is in line with the genetic correlation between autism and schizophrenia. It is unclear if reduction in ICVF is a transdiagnostic marker of neurodevelopmental and mental health conditions, or a marker of a co-occurring condition that is shared between autism and schizophrenia, for example, depression and anxiety.

The association with ICVF was widespread across the cortex and differed regionally between ABCD and the UKB. This may reflect age related changes in ICVF, age related genetic effects, or simply heterogeneity between cohorts. However, in both cohorts, for ICVF there was a strong correlation between hubness and the magnitude of association with autism PGS. One plausible explanation for this is the shared genetics between intracortical connectivity and autism. In other words, the same underlying mechanism that results in greater connectivity among regions may also contribute to variation in autistic traits. Similar findings have also been observed with schizophrenia [30, 72], suggesting broader shared genetics between determinants of hubness and neurodevelopmental and psychiatric outcomes.

In addition to ICVF, this study has also found significant global associations between PGS for autism, CT and SA in the adult study cohort, but not in the ABCD, possibly because of relatively small sample sizes leading to low statistical power to detect small effects. Large-scale case-control neuroimaging studies and meta-analysis have identified increased global CT and grey matter volume in autism [9, 11]. For regional phenotypes, beyond ICVF, this study observed significant associations between PGS for autism and SA for adults, associations between PGS for autism and MC for children. The lack of consistent findings across cohorts suggests that other factors, such as age difference between cohorts might also modify the association between autism and brain structure. The validity of these findings will require further testing using additional datasets and methods. Additionally, we also did not replicate previous regional findings in CT [26], potentially due to methodological differences or due to regression towards the mean.

The lack of consistent association between PGS for autism and SA, a measure of cortical expansion, is noteworthy. Previous research from our group has demonstrated an enrichment of polygenic signals for SA in or near genes associated with neurodevelopmental conditions (including autism) from studies of *de novo* and rare variants [22]. Furthermore, both neurodevelopmental genes and common genetic variants for SA are enriched for neural progenitor cells [22]. Neither of these observations are true for ICVF. Although both common and rare genetic variants are associated with autism, questions still remain if these two broad classes of variants lead to the same phenotype in the brain [23, 25] and are enriched in the same biological processes [73]. The differential overlap between rare-genetic variants and common genetic variants for autism with SA and ICVF respectively, suggests, partly different neurological effects.

For sex differences, we observed some sex-specific global associations between sex-stratified PGS and MRI-derived phenotype in sex-stratified sample populations, but no association was observed for regional phenotypes. Furthermore, we did not have any robust evidence to believe that the effect of PGS for autism on structural brain phenotypes, either globally or regionally differs by sex. The current results provide no evidence to suggest that sex differences in autism diagnosis and presentation emerge from sex differences in brain structure. However, we note that the sex-stratified GWAs were not particularly well powered, and that the sex-stratified analyses were conducted using smaller sample sizes compared to the unstratified analysis.

Despite the large sample size, our study has a few limitations. First, we focussed only on individuals of genetically inferred European ancestries given both the relatively small number of individuals of other ancestries with neuroimaging and genetic data, and the poor portability of polygenic scores across genetically inferred ancestries [74]. However, we have no reason to believe that these results will be discrepant in other genetically inferred ancestry groups. Second, our study does not interrogate how heterogeneity within autism [23] may contribute to differences in results. Third, the current polygenic scores for autism capture only between 1 - 2% of the total variance for autism [3, 23]. However significant findings from polygenic scores have been corroborated using genetic correlations, which accounts for a larger fraction of the variance in autism likelihood.

In conclusion, we find robust evidence to suggest that common variants that increase the likelihood for autism are associated with decreased neurite density index. This supports similar findings from smaller-scale neuroimaging, animal and postmortem research. However, this association with neurite density is not specific to autism, and future research needs to investigate whether reduced neurite density is a transdiagnostic marker for neurodevelopmental and mental health conditions.

## Data Availability

All raw code and data produced are available online at https://github.com/yg330/autism.img-pgs. GWAS summary statistics and cortical morphology data are available through request from the CAM:IDE Data Access Portal (https://portal.ide-cam.org.uk/overview/483). iPSYCH-based GWAS data for autism [49] can be requested from Jakob Grove and Anders Borglum.

https://github.com/ucam-department-of-psychiatry/maps_and_parcs

## Acknowledgements

This research was supported by funding from the Simons Foundation for Autism Research Initiative, the Wellcome Trust (214322\Z\18\Z), Horizon-Europe R2D2-MH (grant agreement number 101057385), and UKRI (10063472). For the purpose of open access, we have applied a CC BY public copyright licence to any author-accepted manuscript version arising from this submission. S.B.-C. also received funding from the Autism Centre of Excellence, the Templeton World Charitable Fund, the MRC and the National Institute for Health Research Cambridge Biomedical Research Centre. The research was supported by the National Institute for Health Research Applied Research Collaboration East of England. Any views expressed are those of the author(s) and not necessarily those of the funder. Some of the results leading to this publication have received funding from the Innovative Medicines Initiative 2 Joint Undertaking under grant agreement no. 777394 for the project AIMS-2-TRIALS. This joint undertaking receives support from the European Union’s Horizon 2020 research and innovation program and the EFPIA and Autism Speaks, Autistica and the SFARI. The iPSYCH team was supported by grants from the Lundbeck Foundation (R102-A9118, R155-2014-1724 and R248-2017-2003), the NIMH (1R01MH124851-01 to A.D.B.), and EU’s Horizon Europe program (R2D2-MH; grant agreement no. 101057385 to A.D.B.). The Danish National Biobank resource was supported by the Novo Nordisk Foundation. High-performance computer capacity for handling and statistical analysis of iPSYCH data on the GenomeDK HPC facility was provided by the Center for Genomics and Personalized Medicine and the Centre for Integrative Sequencing, iSEQ, Aarhus University, Denmark (grant to A.D.B.).

## Ethics declarations

The authors declare no competing interests.

## APEX Consortium

Deep Adhya, Carrie Allison, Bonnie Ayeung, Rosie Bamford, Simon Baron-Cohen, Richard Bethlehem, Tal Biron-Shental, Graham Burton, Wendy Cowell, Jonathan Davies, Joanna Davis, Dori Floris, Alice Franklin, Lidia Gabis, Daniel Geschwind, David M. Greenberg, Yuanjun Gu, Alexandra Havdahl, Alexander Heazell, Rosemary Holt, Matthew Hurles, Yumnah Khan, Meng-Chuan Lai, Madeline Lancaster, Michael Lombardo, Hilary Martin, Jose Gonzalez Martinez, Jonathan Mill, Mahmoud Musa, Kathy Niakan, Adam Pavlinek, Lucia Dutan Polit, Marcin Radecki, David Rowitch, Jenifer Sakai, Laura Sichlinger, Deepak Srivastava, Alexandros Tsompanidis, Florina Uzefovsky, Varun Warrier, Elizabeth Weir, Xinhe Zhang.

## iPSYCH Autism working group

Anders Borglum, Jonas Bybjerg-Grauholm, Jakob Grove, David M. Hougaard, Ole Mors, Preben Bo Mortensen, Merete Nordentoft and Thomas Werge.

## Notes

### Competing Interest Statement

The authors have declared no competing interest.

### Funding Statement

This study was funded by the Simons Foundation; Wellcome Trust (Wellcome), ref number 214322\Z\18\Z; EU Framework Programme for Research and Innovation H2020, ref number 101057385; and UK Research and Innovation , Grant Reference Number 10063472

### Author Declarations

This study is based on data from two independent cohorts, the UK Biobank (UKB) and the Adolescent Brain Cognitive Development (ABCD) database. Ethical approval was obtained from the Human Biology Research Ethics Committee, University of Cambridge (Cambridge, UK). Informed consent was provided by all participants.

## References

1. Tick B, Bolton P, Happé F, Rutter M, Rijsdijk F. Heritability of autism spectrum disorders: a meta-analysis of twin studies. J Child Psychol Psychiatry. 2016;57:585–595.

2. Bai D, Yip BHK, Windham GC, Sourander A, Francis R, Yoffe R, et al. Association of Genetic and Environmental Factors With Autism in a 5-Country Cohort. JAMA Psychiatry. 2019;76:1035–1043.

3. Grove J, Ripke S, Als TD, Mattheisen M, Walters RK, Won H, et al. Identification of common genetic risk variants for autism spectrum disorder. Nat Genet. 2019;51:431– 444.

4. Gaugler T, Klei LL, Sanders SJ, Bodea CA, Goldberg AP, Lee AB, et al. Most genetic risk for autism resides with common variation. Nat Genet. 2014;46:881–885.

5. Parikshak NN, Swarup V, Belgard TG, Irimia M, Ramaswami G, Gandal MJ, et al. Genome-wide changes in lncRNA, splicing, and regional gene expression patterns in autism. Nature. 2016;540:423–427.

6. Satterstrom FK, Kosmicki JA, Wang J, Breen MS, De Rubeis S, An J-Y, et al. Large-Scale Exome Sequencing Study Implicates Both Developmental and Functional Changes in the Neurobiology of Autism. Cell. 2020;180:568–584.e23.

7. Romero-Garcia R, Warrier V, Bullmore ET, Baron-Cohen S, Bethlehem RAI. Synaptic and transcriptionally downregulated genes are associated with cortical thickness differences in autism. Mol Psychiatry. 2018. 2018. 10.1038/s41380-018-0023-7.

8. Gandal MJ, Haney JR, Wamsley B, Yap CX, Parhami S, Emani PS, et al. Broad transcriptomic dysregulation occurs across the cerebral cortex in ASD. Nature. 2022;611:532–539.

9. Bethlehem RAI, Seidlitz J, White SR, Vogel JW, Anderson KM, Adamson C, et al. Brain charts for the human lifespan. Nature. 2022;604:525–533.

10. van Rooij D, Anagnostou E, Arango C, Auzias G, Behrmann M, Busatto GF, et al. Cortical and Subcortical Brain Morphometry Differences Between Patients With Autism Spectrum Disorder and Healthy Individuals Across the Lifespan: Results From the ENIGMA ASD Working Group. Am J Psychiatry. 2018;175:359–369.

11. Bedford SA, Park MTM, Devenyi GA, Tullo S, Germann J, Patel R, et al. Large-scale analyses of the relationship between sex, age and intelligence quotient heterogeneity and cortical morphometry in autism spectrum disorder. Mol Psychiatry. 2020;25:614–628.

12. Buch AM, Vértes PE, Seidlitz J, Kim SH, Grosenick L, Liston C. Molecular and network-level mechanisms explaining individual differences in autism spectrum disorder. Nat Neurosci. 2023;26:650–663.

13. Shan X, Uddin LQ, Xiao J, He C, Ling Z, Li L, et al. Mapping the Heterogeneous Brain Structural Phenotype of Autism Spectrum Disorder Using the Normative Model. Biol Psychiatry. 2022;91:967–976.

14. Hwang G, Wen J, Sotardi S, Brodkin ES, Chand GB, Dwyer DB, et al. Three imaging endophenotypes characterize neuroanatomical heterogeneity of autism spectrum disorder. bioRxiv. 2022.

15. Pretzsch CM, Schäfer T, Lombardo MV, Warrier V, Mann C, Bletsch A, et al. Neurobiological Correlates of Change in Adaptive Behavior in Autism. Am J Psychiatry. 2022;179:336–349.

16. Bethlehem RAI, Seidlitz J, Romero-Garcia R, Trakoshis S, Dumas G, Lombardo MV. A normative modelling approach reveals age-atypical cortical thickness in a subgroup of males with autism spectrum disorder. Commun Biol. 2020;3:486.

17. Lombardo MV, Eyler L, Pramparo T, Gazestani VH, Hagler DJ Jr, Chen C-H, et al. Atypical genomic cortical patterning in autism with poor early language outcome. Sci Adv. 2021;7:eabh1663.

18. Alemany S, Blok E, Jansen PR, Muetzel RL, White T. Brain morphology, autistic traits, and polygenic risk for autism: A population-based neuroimaging study. Autism Res. 2021;14:2085–2099.

19. Arunachalam Chandran V, Pliatsikas C, Neufeld J, O’Connell G, Haffey A, DeLuca V, et al. Brain structural correlates of autistic traits across the diagnostic divide: A grey matter and white matter microstructure study. Neuroimage Clin. 2021;32:102897.

20. Hazlett HC, Gu H, Munsell BC, Kim SH, Styner M, Wolff JJ, et al. Early brain development in infants at high risk for autism spectrum disorder. Nature. 2017;542:348– 351.

21. Hazlett HC, Poe M, Gerig G, Styner M, Chappell C, Smith RG, et al. Early Brain Overgrowth in Autism Associated with an Increase in Cortical Surface Area Before Age 2 years. Arch Gen Psychiatry. 2011;68:467–476.

22. Warrier V, Stauffer E-M, Huang QQ, Wigdor EM, Slob EAW, Seidlitz J, et al. Genetic insights into human cortical organization and development through genome-wide analyses of 2,347 neuroimaging phenotypes. Nat Genet. 2023;55:1483–1493.

23. Warrier V, Zhang X, Reed P, Havdahl A, Moore TM, Cliquet F, et al. Genetic correlates of phenotypic heterogeneity in autism. Nat Genet. 2022. 2 June 2022. 10.1038/s41588-022-01072-5.

24. Weiner DJ, Wigdor EM, Ripke S, Walters RK, Kosmicki JA, Grove J, et al. Polygenic transmission disequilibrium confirms that common and rare variation act additively to create risk for autism spectrum disorders. Nat Genet. 2017;49:978–985.

25. Rolland T, Cliquet F, Anney RJL, Moreau C, Traut N, Mathieu A, et al. Phenotypic effects of genetic variants associated with autism. Nat Med. 2023;29:1671–1680.

26. Khundrakpam B, Vainik U, Gong J, Al-Sharif N, Bhutani N, Kiar G, et al. Neural correlates of polygenic risk score for autism spectrum disorders in general population. Brain Commun. 2020;2:fcaa092.

27. Sha Z, Schijven D, Francks C. Patterns of brain asymmetry associated with polygenic risks for autism and schizophrenia implicate language and executive functions but not brain masculinization. Mol Psychiatry. 2021;26:7652–7660.

28. Shen X, Howard DM, Adams MJ, Hill WD, Clarke T-K, Major Depressive Disorder Working Group of the Psychiatric Genomics Consortium, et al. A phenome-wide association and Mendelian Randomisation study of polygenic risk for depression in UK Biobank. Nat Commun. 2020;11:2301.

29. Stauffer E-M, Bethlehem RAI, Warrier V, Murray GK, Romero-Garcia R, Seidlitz J, et al. Grey and white matter microstructure is associated with polygenic risk for schizophrenia. Mol Psychiatry. 2021;26:7709–7718.

30. Stauffer E-M, Bethlehem RAI, Dorfschmidt L, Won H, Warrier V, Bullmore ET. The genetic relationships between brain structure and schizophrenia. Nat Commun. 2023;14:7820.

31. Floris DL, Peng H, Warrier V, Lombardo MV, Pretzsch CM, Moreau C, et al. The Link Between Autism and Sex-Related Neuroanatomy, and Associated Cognition and Gene Expression. Am J Psychiatry. 2023;180:50–64.

32. Ritchie SJ, Cox SR, Shen X, Lombardo MV, Reus LM, Alloza C, et al. Sex Differences in the Adult Human Brain: Evidence from 5216 UK Biobank Participants. Cereb Cortex. 2018;28:2959–2975.

33. Wierenga LM, Doucet GE, Dima D, Agartz I, Aghajani M, Akudjedu TN, et al. Greater male than female variability in regional brain structure across the lifespan. Hum Brain Mapp. 2022;43:470–499.

34. Williams CM, Peyre H, Toro R, Ramus F. Neuroanatomical norms in the UK Biobank: The impact of allometric scaling, sex, and age. Hum Brain Mapp. 2021;42:4623–4642.

35. Lai M-C, Szatmari P. Sex and gender impacts on the behavioural presentation and recognition of autism. Curr Opin Psychiatry. 2020;33:117–123.

36. Loomes R, Hull L, Mandy WPL. What Is the Male-to-Female Ratio in Autism Spectrum Disorder? A Systematic Review and Meta-Analysis. J Am Acad Child Adolesc Psychiatry. 2017;56:466–474.

37. Yeo BTT, Krienen FM, Sepulcre J, Sabuncu MR, Lashkari D, Hollinshead M, et al. The organization of the human cerebral cortex estimated by intrinsic functional connectivity. J Neurophysiol. 2011;106:1125–1165.

38. Mesulam MM. From sensation to cognition. Brain. 1998;121 ( Pt 6):1013–1052.

39. Sudlow C, Gallacher J, Allen N, Beral V, Burton P, Danesh J, et al. UK Biobank: An Open Access Resource for Identifying the Causes of a Wide Range of Complex Diseases of Middle and Old Age. PLoS Med. 2015;12:e1001779.

40. Casey BJ, Cannonier T, Conley MI, Cohen AO, Barch DM, Heitzeg MM, et al. The Adolescent Brain Cognitive Development (ABCD) study: Imaging acquisition across 21 sites. Dev Cogn Neurosci. 2018. 14 March 2018. 10.1016/J.DCN.2018.03.001.

41. Fischl B, van der Kouwe A, Destrieux C, Halgren E, Ségonne F, Salat DH, et al. Automatically parcellating the human cerebral cortex. Cereb Cortex. 2004;14:11–22.

42. Glasser MF, Coalson TS, Robinson EC, Hacker CD, Harwell J, Yacoub E, et al. A multi-modal parcellation of human cerebral cortex. Nature. 2016;536:171–178.

43. Rosen AFG, Roalf DR, Ruparel K, Blake J, Seelaus K, Villa LP, et al. Quantitative assessment of structural image quality. Neuroimage. 2018;169:407–418.

44. Daducci A, Canales-Rodríguez EJ, Zhang H, Dyrby TB, Alexander DC, Thiran J-P. Accelerated Microstructure Imaging via Convex Optimization (AMICO) from diffusion MRI data. Neuroimage. 2015;105:32–44.

45. de Groot M, Vernooij MW, Klein S, Ikram MA, Vos FM, Smith SM, et al. Improving alignment in Tract-based spatial statistics: evaluation and optimization of image registration. Neuroimage. 2013;76:400–411.

46. Bycroft C, Freeman C, Petkova D, Band G, Elliott LT, Sharp K, et al. The UK Biobank resource with deep phenotyping and genomic data. Nature. 2018;562:203–209.

47. Warrier V, Kwong ASF, Luo M, Dalvie S, Croft J, Sallis HM, et al. Gene–environment correlations and causal effects of childhood maltreatment on physical and mental health: a genetically informed approach. The Lancet Psychiatry. 2021;8:373–386.

48. Ge T, Chen C-Y, Ni Y, Feng Y-CA, Smoller JW. Polygenic prediction via Bayesian regression and continuous shrinkage priors. Nat Commun. 2019;10:1776.

49. Bybjerg-Grauholm J, Bøcker Pedersen C, Bækvad-Hansen M, Giørtz Pedersen M, Adamsen D, Søholm Hansen C, et al. The iPSYCH2015 Case-Cohort sample: updated directions for unravelling genetic and environmental architectures of severe mental disorders. bioRxiv. 2020.

50. Benjamini Y, Hochberg Y. Controlling the false discovery rate: A practical and powerful approach to multiple testing. J R Stat Soc. 1995;57:289–300.

51. Bulik-Sullivan B, Finucane HK, Anttila V, Gusev A, Day FR, Loh P-R, et al. An atlas of genetic correlations across human diseases and traits. Nat Genet. 2015;47:1236–1241.

52. 1000 Genomes Project Consortium, Auton A, Brooks LD, Durbin RM, Garrison EP, Kang HM, et al. A global reference for human genetic variation. Nature. 2015;526:68– 74.

53. Alexander-Bloch AF, Shou H, Liu S, Satterthwaite TD, Glahn DC, Shinohara RT, et al. On testing for spatial correspondence between maps of human brain structure and function. Neuroimage. 2018;178:540–551.

54. Jiang L, Zheng Z, Qi T, Kemper KE, Wray NR, Visscher PM, et al. A resource-efficient tool for mixed model association analysis of large-scale data. Nat Genet. 2019;51:1749– 1755.

55. Hemani G, Zheng J, Elsworth B, Wade KH, Haberland V, Baird D, et al. The MR-Base platform supports systematic causal inference across the human phenome. Elife. 2018;7.

56. Hemani G, Tilling K, Davey Smith G. Orienting the causal relationship between imprecisely measured traits using GWAS summary data. PLoS Genet. 2017;13:e1007081.

57. Verbanck M, Chen C-Y, Neale B, Do R. Detection of widespread horizontal pleiotropy in causal relationships inferred from Mendelian randomization between complex traits and diseases. Nat Genet. 2018;50:693–698.

58. Khundrakpam B, Bhutani N, Vainik U, Gong J, Al-Sharif N, Dagher A, et al. A critical role of brain network architecture in a continuum model of autism spectrum disorders spanning from healthy individuals with genetic liability to individuals with ASD. Mol Psychiatry. 2023;28:1210–1218.

59. Grussu F, Schneider T, Tur C, Yates RL, Tachrount M, Ianuş A, et al. Neurite dispersion: a new marker of multiple sclerosis spinal cord pathology? Ann Clin Transl Neurol. 2017;4:663–679.

60. Gong N-J, Dibb R, Pletnikov M, Benner E, Liu C. Imaging microstructure with diffusion and susceptibility MR: neuronal density correlation in Disrupted-in-Schizophrenia-1 mutant mice. NMR Biomed. 2020;33:e4365.

61. Genc S, Malpas CB, Holland SK, Beare R, Silk TJ. Neurite density index is sensitive to age related differences in the developing brain. Neuroimage. 2017;148:373–380.

62. Slater DA, Melie-Garcia L, Preisig M, Kherif F, Lutti A, Draganski B. Evolution of white matter tract microstructure across the life span. Hum Brain Mapp. 2019;40:2252– 2268.

63. Andica C, Kamagata K, Kirino E, Uchida W, Irie R, Murata S, et al. Neurite orientation dispersion and density imaging reveals white matter microstructural alterations in adults with autism. Mol Autism. 2021;12:48.

64. Arai T, Kamagata K, Uchida W, Andica C, Takabayashi K, Saito Y, et al. Reduced neurite density index in the prefrontal cortex of adults with autism assessed using neurite orientation dispersion and density imaging. Front Neurol. 2023;14:1110883.

65. Carper RA, Treiber JM, White NS, Kohli JS, Müller R-A. Restriction Spectrum Imaging As a Potential Measure of Cortical Neurite Density in Autism. Front Neurosci. 2016;10:610.

66. Barnett BR, Torres-Velázquez M, Yi SY, Rowley PA, Sawin EA, Rubinstein CD, et al. Sex-specific deficits in neurite density and white matter integrity are associated with targeted disruption of exon 2 of the Disc1 gene in the rat. Transl Psychiatry. 2019;9:82.

67. Lee E, Lee J, Kim E. Excitation/Inhibition Imbalance in Animal Models of Autism Spectrum Disorders. Biol Psychiatry. 2017;81:838–847.

68. Stoner R, Chow ML, Boyle MP, Sunkin SM, Mouton PR, Roy S, et al. Patches of disorganization in the neocortex of children with autism. N Engl J Med. 2014;370:1209– 1219.

69. Casanova MF, Buxhoeveden DP, Switala AE, Roy E. Minicolumnar pathology in autism. Neurology. 2002;58:428–432.

70. Kamiya K, Hori M, Aoki S. NODDI in clinical research. J Neurosci Methods. 2020;346:108908.

71. Kraguljac NV, Guerreri M, Strickland MJ, Zhang H. Neurite Orientation Dispersion and Density Imaging in Psychiatric Disorders: A Systematic Literature Review and a Technical Note. Biological Psychiatry Global Open Science. 2023;3:10.

72. Morgan SE, Seidlitz J, Whitaker KJ, Romero-Garcia R, Clifton NE, Scarpazza C, et al. Cortical patterning of abnormal morphometric similarity in psychosis is associated with brain expression of schizophrenia-related genes. Proc Natl Acad Sci U S A. 2019;116:9604–9609.

73. Weiner DJ, Ling E, Erdin S, Tai DJC, Yadav R, Grove J, et al. Statistical and functional convergence of common and rare genetic influences on autism at chromosome 16p. Nat Genet. 2022;54:1630–1639.

74. Ding Y, Hou K, Xu Z, Pimplaskar A, Petter E, Boulier K, et al. Polygenic scoring accuracy varies across the genetic ancestry continuum. Nature. 2023. 17 May 2023. 10.1038/s41586-023-06079-4.

