## Supplementary Figures for "Polygenic scores for autism are associated with neurite density in adults and children from the general population"

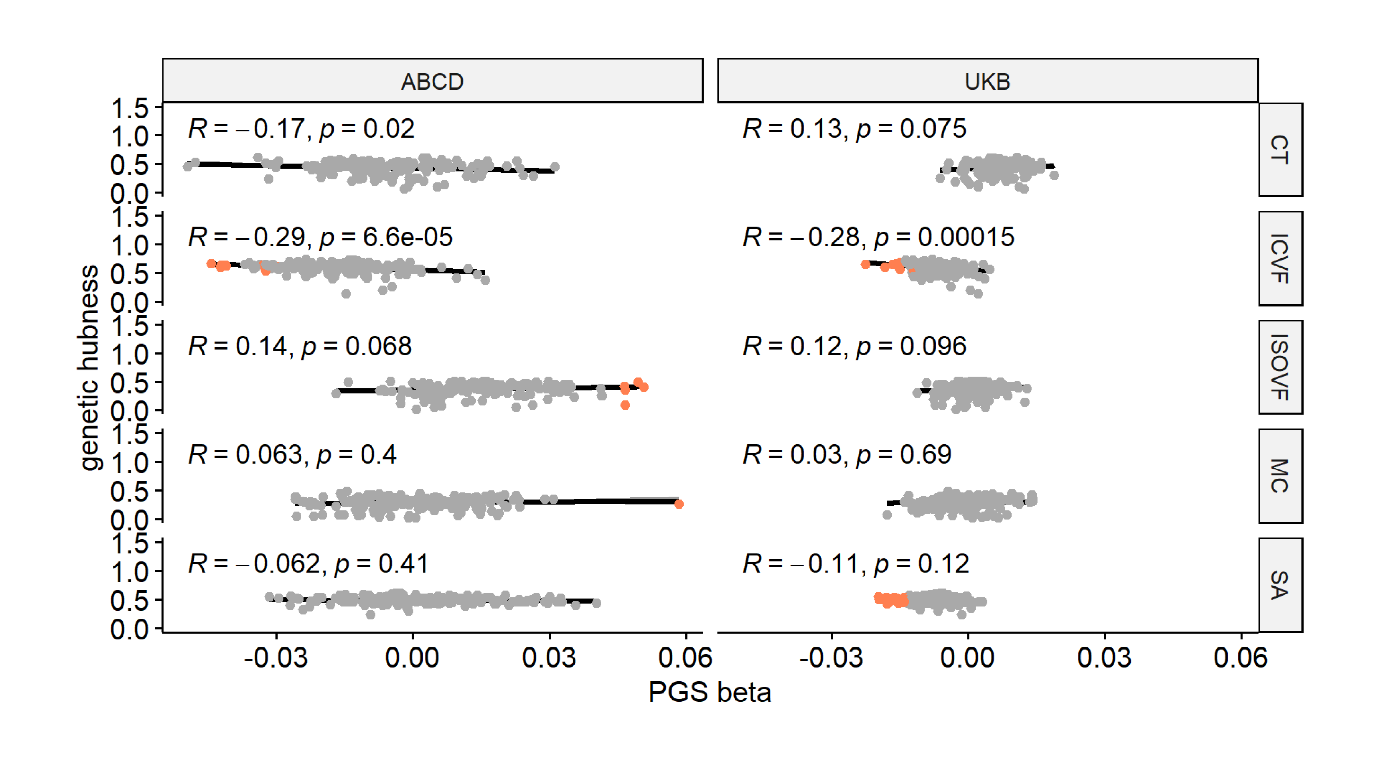

**Supplementary Figure 1. Enrichment Analysis:** *Correlation plot between genetic hubness and autism PGS association across MRI-derived phenotypes of interest in the ABCD and UK Biobank cohorts. Each point represents a region and regions where there is a significant association between the phenotype and autism PGS are coloured in red. R is the Pearson correlation coefficient. p is the p value before multiple testing correction.*

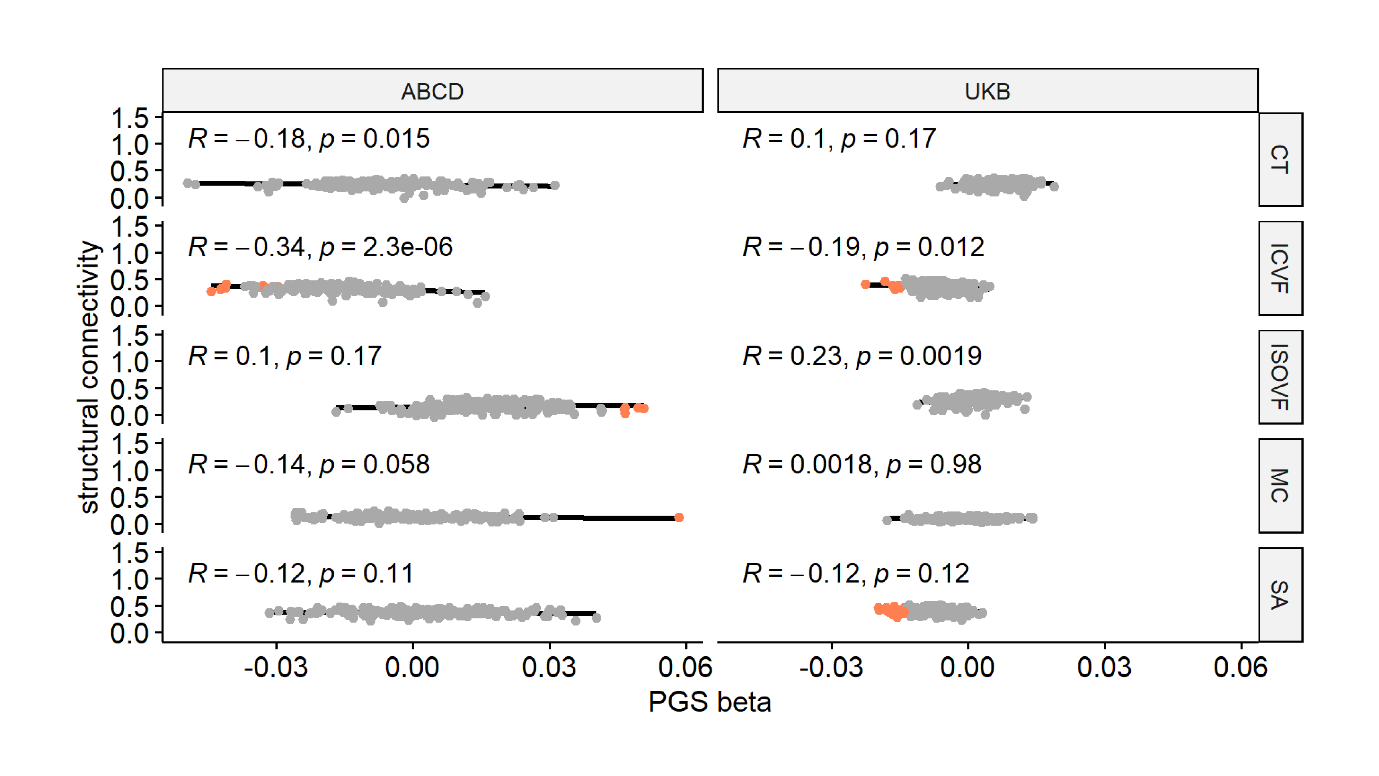

**Supplementary Figure 2. Enrichment Analysis:** *Correlation plot between structural connectivity and autism PGS association across MRI-derived phenotypes of interest in the ABCD and UK Biobank cohorts. Each point represents a region and regions where there is a significant association between the phenotype and autism PGS are coloured in red. R is the Pearson correlation coefficient. p is the p value before multiple testing correction.*
